# An exploratory factor analysis approach to identifying patient benefits of a cost-of-care conversation tool in routine prenatal care

**DOI:** 10.1101/2023.03.02.23286699

**Authors:** Anne Rivelli, Veronica Fitzpatrick, Maureen Shields, Kim Erwin

**Author notes:** Corresponding Author: Veronica Fitzpatrick, DrPH, MPH.

## Abstract

To address the need for cost-of-care conversations in prenatal care, the CONTINUE (**c**ost c**on**versa**ti**ons i**n** ro**u**tin**e** prenatal care) study was conducted with prenatal patients to better understand the benefits of implementing a cost-of-care conversation “cost” tool into routine obstetrics (OB) care. This research team conducted a multi-phase, mixed-methods research study to identify 18 target benefits of a cost tool to initiate and standardize cost-of-care conversation and, subsequently, developed and validated a cost tool. The cost tool was piloted and data pertaining to cost tool benefits were collected through interviews and surveys. To comprehensively assess the cost tool’s utility, exploratory factor analysis was performed to classify the underlying factor structure of the 18 benefit item responses. Data includes patients’ self-reported experiences of benefit items, as collected from third trimester prenatal patient participants who received the tool at the beginning of their prenatal care in three midwestern-based hospital clinics within one healthcare system. The present study describes the factor analysis approach used to identify the three final factors that emerged from the data. This analysis provides a framework for exploring patient-specific predictors of experiencing the benefit-related factors of a cost tool incorporated into routine OB care.

**Statements and Declarations:** This study was funded by the Robert Wood Johnson Foundation, #77290.

## Introduction

Although there are many reasons why patients miss prenatal appointments, research indicates that some groups of women are more likely to forgo prenatal care because of logistical and issues like hourly jobs, transportation, lack of childcare, and inadequate social support (Kaiser Family Foundation 2018). Cost-of-care conversations, or conversations between patients and providers about logistical and cost-related factors that exist for patients, could help mitigate barriers to prenatal care. Research has shown that cost-of-care conversations can lead to better health outcomes by improving patient adherence to treatment plans and enhancing patient engagement in the management of their care (Meluch & Oglesby 2015; Piette, Heisler, Wagner 2004). Because prenatal care visits are typically scheduled to be 10-15 minutes each and biomedical needs are often prioritized in these visits, conversations about unmet patient needs and other cost-related barriers to care are not frequently initiated (Rising 1998; Novick 2009). The use of tools to supplement otherwise routine prenatal care have shown promise, particularly to promote communication between patients and providers (Ngo, Truong, Nordeng 2020).

To address the need for cost-of-care conversations in prenatal care, a multi-phased, mixed-methods pilot study (“CONTINUE”, or cost conversations in routine prenatal care) was conducted with pregnant patients to identify the benefits of implementing a cost tool into routine obstetrics (OB) care. In the first phase of the CONTINUE study, a cost tool was created to address the need for a means to initiate and standardize cost-of-care conversations in prenatal care. The tool was created to be personalized to the patient, reflecting the prenatal care plan decided between the patient and provider at the onset of prenatal care. The cost tool’s purpose is to provide patients with a visual forecasting of their remaining prenatal care, including expected appointments, labs, scans, and tests, as well as estimated time required for each visit and other help resources that help patients navigate their care. The tool was designed to benefit patients; therefore, during the first phase of the CONTINUE study, 18 potential benefits of the cost tool (“target benefit items”) were also hypothesized (Erwin et al. 2019).

In the second and final phase of the CONTINUE study, the cost tool was tailored to three different OB settings and prenatal patients were interviewed to validate the target benefit items as true potential benefits of the cost tool. After target benefit items were validated, the cost tool was incorporated into routine OB care in the same three clinics serving a diverse patient population. Patients were given the cost tool to use throughout their prenatal care and, at the end of their prenatal care, patients provided responses rating their agreement with each of the 18 benefits items they experienced from the use of the cost tool.

The purpose of this paper is two-fold. First, this paper will describe a method for unifying 18 benefit item responses derived from two different data collection methods, on different measurement scales, in a mixed-methods study. Second, this paper will reveal key constructs underlying the 18 benefits items for theme and brevity. This team utilized factor analysis to identify common underlying factors that can explain interrelationships between the 18 benefit items. The identification of factors derived from this analysis will allow researchers a framework for exploring these factors as outcomes while preserving all original benefits within the common factor. Furthermore, identification of these factors will establish the main benefits that have been demonstrated among prenatal patients due to the use of a cost tool. This has implications for widespread use of this tool, and others like it, to bridge the gap in cost-of-care conversations between prenatal patients and their providers.

## Methods

The CONTINUE study is a multi-phased observational pilot study that documented the effects of implementing a validated cost tool into routine care in three OB clinics in a midwestern-based healthcare system. Site providers were trained on how to personalize and share the cost tool with prenatal patients. The cost tool was available in both English and Spanish and offered to patients in their preferred language. Providers were encouraged to share the tool to all early-stage prenatal patients but were given discretion regarding with whom it was given. Tool implementation occurred between September 2020 and July 2021, after study approval was granted by the health system’s institutional review board (#20-264E).

At the end of tool implementation, prenatal patients who received the cost tool were passively recruited to participate, meaning flyers and posters with study contact info in both English and Spanish were displayed in clinic waiting rooms and clinic rooms for patients to initiate participation in either an in-person or virtual semi-structured interview or electronic survey to provide feedback on their experiences with the cost tool.

Prenatal patient participants were at least 18 years old, at least 27 weeks pregnant, and had received the tool at least 8 weeks prior to being interviewed or surveyed. Participants self-selected their participation method, as interview (in-person at a public location of the patient’s choosing or virtual) or electronic survey, as well as their preferred language (English or Spanish). Surveys were provided and interviews were conducted in the preferred language.

Patient interviews and surveys were conducted to assess uptake, use, and benefits of the tool. Benefit items were originally determined from patient interviews using a card sort method validated in the field of human-centered design (HCD) (Chen, Neta and Roberts 2021). In the case of this study, participants were given cards with each of the 18 benefit items and asked to sort each as benefits they experienced, did not experience, or neither (i.e. neutral) from the use of the tool. Broadly, per Chen, Neta and Roberts (2021), the card sort method engages patients in the implementation effort and leads to better tailored and more adaptable products – in this study, a cost tool. The CONTINUE study utilized the 18 benefit items as outcomes of interest to understand experiences of the cost tool among patients. Some examples of benefit items include: “The pregnancy planning guide helped me navigate insurance more effectively”, “The pregnancy planning guide helped me feel my financial situation was being considered.”. “The pregnancy planning guide helped me show up on time.” Observed data was collected from patient interviews and surveys and unified for analysis. Now, factor analysis is being applied to identify common factors underlying tool benefit items.

Factor analysis, in general, refers to all methods of data analysis that use matrix factors. It is a technique used to reduce a large number of items into a fewer number of factors, or constructs encompassing shared underlying characteristics of the original, observed items used to create them (Child 1990). The basic assumption of factor analysis is that, for a collection of observed items, there are underlying variables called factors that can explain the interrelationships among those items. Therefore, the aim of factor analysis is to reveal any latent factors that cause the observed items to covary, thereby reducing the observed items into a smaller number of common factors (Costello & Osborne 2005). It’s a flexible approach to a pragmatic method of analysis. Exploratory factor analysis (EFA) was applied in this analysis, with a goal to identify factors to serve as outcomes in follow-up pilot study analyses.

### Measures and Data

Data (i.e. observed item responses) come from a pilot study assessing potential benefits of a pregnancy support tool implemented to encourage cost conversations between patients receiving prenatal care and their providers. The study included 71 prenatal patient participants receiving care from one of three OB clinics within a single, large midwestern-based healthcare system who had received the cost tool from their provider early in their prenatal care. The cost tool was available in both English or Spanish, and patients self-selected which version they preferred to receive. Demographically, participants were primarily ages 26-35 (57.75%), White (71.43%), and Hispanic/Latino/Spanish (56.34%). Medically, participants were evenly split in insurance, risk status, parity; specifically, 56.34% were publicly insured during the pregnancy, 54.93% experienced low risk pregnancies, and 56.34% were experiencing subsequent pregnancies (56.34%). Finally, data were self-reported by patient participants via two different data collection methods, including interviews (52.11%) and surveys (47.89%). See Figure 1 for patient sample demographics.

**Figure I.**
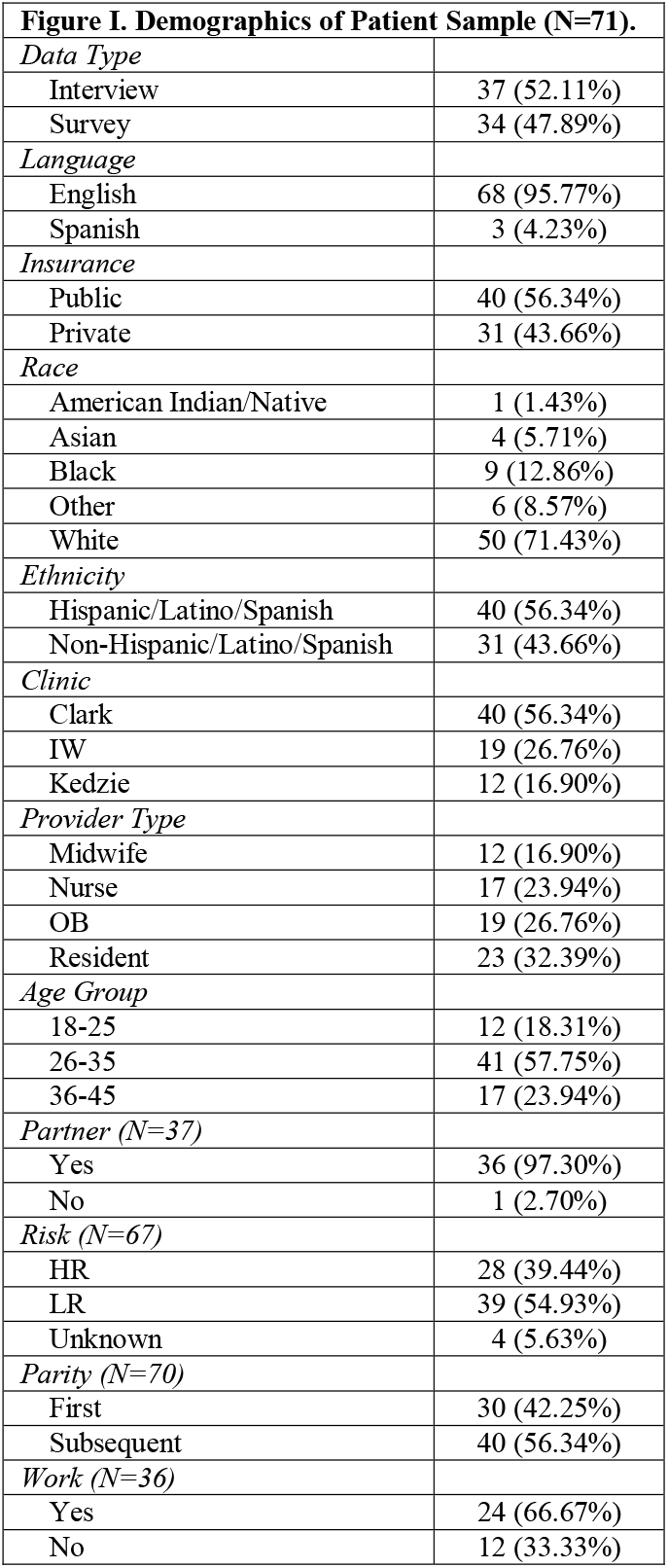
Demographics of Patient Sample (N=71).

Benefit items were assessed using one of the two data collection methods, as self-selected by participants. Because benefit items were derived from patient interviews using a card sort method during the initial phase of this mixed-methods study (Erwin et al. 2019), the team captured benefit item experiences using the same method in the second and final phase. Consequently, the first method of data collection was a 60-minute interview. During the card sort activity within the interview, interview participants were asked to group target benefits (each listed on an individual card) into one of three categories: “Experienced”, “Neutral”, “Did not experience”. The second method of data collection was a 20-minute, 35-item self-report electronic survey. Four research team members agreed on the translation of all items from the card sort method into the 18 survey items. Survey participants were asked to indicate how much they experienced (or did not experience) the target benefits by rating each item as “Strongly Agree”, “Agree”, “Neutral”, “Disagree”, “Strongly Disagree”.

For analysis purposes, benefit item responses from interviews and surveys were merged into one unified Likert-level system with three levels, resulting in survey data responses being collapsed into the three categories aligning with interviews: Experienced (i.e. Strongly Agree and Agree), Neutral (i.e. Neutral) and Did not experience (i.e. Strongly Disagree and Disagree). Benefit item values were assigned numerical ratings (0=Did not experience, 1=Neutral, 2=Experienced) which were then used as the observed indicator values in EFA models to determine factor loadings.

### Data Analysis

Data management and analysis were performed by the study research team and conducted using SAS statistical software (Version 9.4; SAS Institute, Cary, NC). The 18 benefit items were used as observed indicators in models. In the absence of data on the underlying structure of these constructs, the goal was the most flexible approach; therefore, EFA was decided a prior as the best approach. Factor analysis (proc factor) was performed on responses to the 18 benefit items. The research team assumed the presence of common, unique and error variance in item responses, which is best accounted for by factor analysis (O’Rourke & Hatcher 2022; Korstanje 2020). Furthermore, the priority of this analysis was to achieve the most useful interpretations to newly-defined dimensions under an assumption of common latent constructs (Korstanje 2020).

Maximum likelihood estimation (MLE; method=ml) was used to extract factors and was followed by an oblique rotation (rotation=promax). In the interest of flexibility, oblique rotation was chosen, as it allows factors to be correlated with one another and accounts for relationships between factors before determining an item’s relationship to the factor (Beavers et al. 2013; “Factor Analysis” 2022). Factor pattern and structure values were reviewed for final assignment of items within factors.

## Results

### Correlations

Correlations between all items were initially explored. All correlations were moderately high, reiterating the appropriateness of oblique rotation (Tabachnick & Fidell 2007). The full correlation matrix among all variables entered into the factor analysis is presented in Table 1. Of the 143 pairwise correlations, 139 correlations were significant at p<0.05 and 60 of the 139 correlations were significant at p<0.0001. Nonsignificant correlations were found between benefit 9 and benefits 13-16. Across all associations, the average absolute association was 0.43, a moderate correlation, suggesting the items were overlapping in construct and factor analysis was suitable.

**Table I.** Intercorrelations between cost tool benefit items.

| **Benefit Items** | **benefit1** | **benefit2** | **benefit3** | **benefit4** | **benefit5** | **benefit6** | **benefit7** | **benefit8** | **benefit9** | **benefit10** | **benefit11** | **benefit12** | **benefit13** | **benefit14** | **benefit15** | **benefit16** | **benefit17** | **benefit18** |
| --- | --- | --- | --- | --- | --- | --- | --- | --- | --- | --- | --- | --- | --- | --- | --- | --- | --- | --- |
| benefit1 | 1 |  |  |  |  |  |  |  |  |  |  |  |  |  |  |  |  |  |
| benefit2 | 0.499** | 1 |  |  |  |  |  |  |  |  |  |  |  |  |  |  |  |  |
| benefit3 | 0.430* | 0.362* | 1 |  |  |  |  |  |  |  |  |  |  |  |  |  |  |  |
| benefit4 | 0.422* | 0.397* | 0.274* | 1 |  |  |  |  |  |  |  |  |  |  |  |  |  |  |
| benefit5 | 0.380* | 0.336* | 0.424* | 0.278* | 1 |  |  |  |  |  |  |  |  |  |  |  |  |  |
| benefit6 | 0.455** | 0.411* | 0.323* | 0.410* | 0.699** | 1 |  |  |  |  |  |  |  |  |  |  |  |  |
| benefit7 | 0.412* | 0.445** | 0.312* | 0.680** | 0.272* | 0.306* | 1 |  |  |  |  |  |  |  |  |  |  |  |
| benefit8 | 0.537** | 0.400* | 0.445** | 0.389* | 0.422* | 0.591** | 0.574** | 1 |  |  |  |  |  |  |  |  |  |  |
| benefit9 | 0.497** | 0.446** | 0.420* | 0.275* | 0.373* | 0.341* | 0.554** | 0.343* | 1 |  |  |  |  |  |  |  |  |  |
| benefit10 | 0.381* | 0.381* | 0.430* | 0.350* | 0.423* | 0.455** | 0.458** | 0.499** | 0.595** | 1 |  |  |  |  |  |  |  |  |
| benefit11 | 0.398* | 0.398* | 0.487** | 0.454** | 0.376* | 0.339* | 0.671** | 0.567** | 0.615** | 0.509** | 1 |  |  |  |  |  |  |  |
| benefit12 | 0.215* | 0.266* | 0.392* | 0.481** | 0.404* | 0.355* | 0.456** | 0.352* | 0.363* | 0.414* | 0.617** | 1 |  |  |  |  |  |  |
| benefit13 | 0.422* | 0.377* | 0.422* | 0.343* | 0.585** | 0.567** | 0.292* | 0.387* | 0.134 | 0.312* | 0.248* | 0.290* | 1 |  |  |  |  |  |
| benefit14 | 0.364* | 0.429* | 0.473** | 0.402* | 0.531** | 0.542** | 0.370* | 0.589** | 0.152 | 0.377* | 0.354* | 0.440* | 0.593** | 1 |  |  |  |  |
| benefit15 | 0.390* | 0.305* | 0.475** | 0.388* | 0.542** | 0.543** | 0.329* | 0.451** | 0.116 | 0.292* | 0.346* | 0.409* | 0.773** | 0.744** | 1 |  |  |  |
| benefit16 | 0.400* | 0.351* | 0.424* | 0.409* | 0.583** | 0.616** | 0.357* | 0.529** | 0.155 | 0.298* | 0.340* | 0.407* | 0.678** | 0.735** | 0.750** | 1 |  |  |
| benefit17 | 0.483** | 0.266* | 0.439* | 0.382* | 0.414* | 0.508** | 0.401* | 0.531** | 0.362* | 0.412* | 0.321* | 0.332* | 0.489** | 0.406* | 0.418* | 0.476** | 1 |  |
| benefit18 | 0.371* | 0.458** | 0.415* | 0.482** | 0.486** | 0.445* | 0.457** | 0.404* | 0.248* | 0.251* | 0.508** | 0.460** | 0.462** | 0.493** | 0.455** | 0.517** | 0.500** | 1 |

\*Statistically significant at $p < 0.05$ .
\*\*Statistically significant at $p < 0.0001$ .

### Model Selection

An initial review of the factor analysis revealed small partial correlations compared to original correlations. Additionally, the Kaiser’s Measure of Sample Adequacy (MSA) was 0.84, suggesting there were enough variables in the analysis to reliably define common factors (“The Factor Procedure” 2022). Finally, the squared multiple correlations (SMCs) were all fairly large, indicating the results of the EFA approach would likely be similar to a PCA approach regardless (Castro-Schilo 2021); regardless, we moved forward with the EFA.

In the first iteration of the factor analysis model, four criteria were used to select the most meaningful number of factors to retain (O’Rourke & Hatcher 2022; Suhr 2022). First, the eigenvalue-one criterion, or the criterion of eigenvalues being greater than 1.00, indicated up to three potential factors. Second, a review of the scree plot revealed substantial breaks at two or three factors, in line with the previous criterion. Third, the proportion of variance was 67.43% with 1 factor, 16.83% with a second factor, 6.52% with a third factor and 4.29% with a fourth factor, again suggesting the ideal final model would include either two or three factors for the most parsimonious model. EFA models with two and three factors were performed and reviewed by the research team. Items that cross-loaded, based on a definition of a regression coefficient within 0.02, were assigned to factors by the research team. Finally, the fourth criterion, interpretability, or interpreting the substantive meaning of the retained factors and verifying it makes sense with what is known about the constructs under investigation, was discussed among the research team to select the number of factors to retain.

The following results detail factor loading and correlation values for three different models: a three-factor model with oblique rotation, a two-factor model with oblique rotation, and a three-factor model with orthogonal rotation. Because the ultimate goals were to preserve all original items and identify the most interpretable model identifying factors that could serve as outcomes for further data analyses, the three-factor model with oblique rotation was selected as the final model and all 18 original benefit items were retained. The additional two models are described in this paper to support and validate of results of the final selected three-factor model.

### EFA Final Model Results

Eight benefit items, 5-6 and 13-18, loaded onto Factor 1 in this factor solution, with loadings ranging from 0.403-0.863 and Factor 1 item correlations ranging from 0.555-0.862. This factor clearly showcased a cost-related factor, which aligned with an a priori conceptualization, and the research team labeled Factor 1 as Logistics.

Five benefit items, 1-3 and 9-10, loaded onto Factor 2, with loadings ranging from 0.334-0.943 and Factor 2 item correlations ranging from 0.515-0.903. Item 3 loaded onto Factors 1 and 2 and, despite coefficients suggesting it load on Factor 1, it was assigned to Factor 2. Given the decision was made a priori to preserve all benefit items within factors, decisions had to be made about where to assign cross-loaded items. In this case, the research team members decided it fit best with Factor 2 based on theoretical relevance. Factor 2 was labeled by the research team as Efficacy.

Finally, five benefit items, 4,8,7, and 11-12, loaded onto Factor 3, with loadings ranging from 0.338-0.873 and Factor 3 item correlations ranging from 0.539-0.911. Benefit 8 loaded onto Factors 1 and 3 but was assigned to Factor 3 based on the coefficients. Factor 3 was labeled [Patient] Understanding. Final communality estimates ranged from 0.355-0.857, indicating each item’s variance was moderately to well-explained by the factors. See Table 2 for the final three-factor model results.

**Table II.** *Final* Exploratory Factor Analysis Model Results with 3-Factor Loading and Oblique (Promax) Rotation.

|  |  |  |  |  |
| --- | --- | --- | --- | --- |
| Benefit Items | The pregnancy planning guide helped me... | Rotated Factor Pattern*<br>(Standardized Regression Coefficients) | Factor Structure**<br>(Correlations) | Final<br>Communality |

|  |  | Factor1 | Factor2 | Factor3 | Factor1:<br>Logistics | Factor2:<br>Efficacy | Factor3:<br>Understanding | Estimates*** |
| --- | --- | --- | --- | --- | --- | --- | --- | --- |
| benefit15 | feel my financial situation was being considered. | *0.863* | -0.112 | 0.089 | **0.862** | 0.260 | 0.431 | 0.753 |
| benefit16 | make the financial tradeoffs needed to get through my pregnancy. | *0.833* | -0.066 | 0.106 | **0.857** | 0.303 | 0.457 | 0.742 |
| benefit13 | navigate insurance more effectively. | *0.826* | -0.015 | -0.019 | **0.812** | 0.288 | 0.355 | 0.659 |
| benefit14 | see the 'Big Picture' and link it to the family budget. | *0.768* | -0.067 | 0.159 | **0.816** | 0.305 | 0.479 | 0.682 |
| benefit5 | attend more appointments. | *0.651* | 0.379 | -0.199 | **0.702** | 0.524 | 0.295 | 0.597 |
| benefit6 | show up on time. | *0.648* | 0.303 | -0.094 | **0.720** | 0.501 | 0.360 | 0.585 |
| benefit18 | manage my other life responsibilities. | *0.414* | 0.054 | 0.327 | **0.585** | 0.377 | 0.545 | 0.441 |
| benefit17 | feel my time was being respected. | *0.403* | 0.264 | 0.113 | **0.555** | 0.474 | 0.433 | 0.398 |
| benefit9 | feel more in control of my pregnancy. | -0.205 | *0.943* | 0.074 | 0.186 | **0.903** | 0.460 | 0.848 |
| benefit10 | feel confident that I was 'doing things right.' | 0.136 | *0.590* | 0.082 | 0.397 | **0.683** | 0.445 | 0.494 |
| benefit1 | increase my trust in the care team. | 0.280 | *0.426* | 0.091 | 0.483 | **0.578** | 0.437 | 0.421 |
| benefit3 | ask questions I wouldn't have thought to ask. | *0.391* | *0.384* | -0.011 | 0.532 | **0.527** | 0.365 | 0.406 |
| benefit2 | be a stronger partner in my care. | 0.208 | *0.334* | 0.201 | 0.427 | **0.515** | 0.467 | 0.355 |
| benefit7 | understand what was coming next in my pregnancy care. | -0.116 | 0.178 | *0.873* | 0.355 | 0.579 | **0.911** | 0.857 |
| benefit4 | understand how appointments differ. | 0.135 | -0.064 | *0.717* | 0.442 | 0.352 | **0.746** | 0.572 |
| benefit11 | plan ahead and feel less stressed. | -0.001 | 0.438 | *0.471* | 0.383 | 0.678 | **0.694** | 0.624 |
| benefit8 | know how to plan for tests. | *0.361* | 0.194 | *0.341* | 0.592 | 0.505 | **0.607** | 0.519 |
| benefit12 | explain my care plan to others (family, employers, other doctors). | 0.227 | 0.189 | *0.338* | 0.454 | 0.447 | **0.539** | 0.369 |
| *Factor pattern displays item loadings. Italics indicate item factor loading. Note: two items loaded onto two factors.<br>**Factor structure displays correlations between individual items and the final two factors. Bolded correlations represent where items were interpreted as loading.<br>***Final communality estimates represent the proportion of each item's variance that can be explained by the factors. |  |  |  |  |  |  |  |  |

Comparing the two-factor model to the three-factor model, the same eight benefits (5-6, 13-18) loaded on Factor 1 in both the two- and three-factor models, with extremely similar item loading and correlation values. Factors 2 and 3 from the three-factor model were collapsed as one single factor in the two-factor model (Factor 2), with loadings ranging from 0.379-0.873 and item correlations ranging from 0.547-0.759. Final communality estimates ranged from 0.387-0.773.

In the three-factor model, item correlations were generally higher, suggesting greater variance accounted for by the separated factors, and communality estimates were larger, indicating a greater proportion of each item’s variance was explained by the separated factors. Furthermore, all factors within the three-factor model had five or more strongly loading items (.50 or better), indicating all three as solid factors (Costello & Osborne 2005). While there is no proven or universally accepted test to select the optimal model (Goldberg 2022), the two-factor model was ultimately deemed less interpretable compared to the three-factor model. See Table 3 for the two-factor model results with oblique (promax) rotation.

**Table III.** Exploratory Factor Analysis Model Results with 2-Factor Loading and Oblique (Promax) Rotation.

|  |  |  |  |  |  |  |
| --- | --- | --- | --- | --- | --- | --- |
| Benefit Items | The pregnancy planning guide helped me... | Rotated Factor Pattern*<br>(Standardized Regression Coefficients) |  | Factor Structure**<br>(Correlations) |  | Final<br>Communality<br>Estimates*** |
|  |  | Factor 1 | Factor 2 | Factor 1:<br>Logistics | Factor 2:<br>Efficacy/Understanding |  |
| benefit15 | feel my financial situation was being considered. | *0.918* | -0.084 | **0.876** | 0.370 | 0.773 |
| benefit16 | make the financial tradeoffs needed to get through my pregnancy. | *0.870* | -0.015 | **0.863** | 0.416 | 0.744 |
| benefit13 | navigate insurance more effectively. | *0.855* | -0.080 | **0.816** | 0.344 | 0.671 |
| benefit14 | see the 'Big Picture' and link it to the family budget. | *0.797* | 0.049 | **0.821** | 0.444 | 0.676 |
| benefit6 | show up on time. | *0.590* | 0.219 | **0.699** | 0.511 | 0.524 |
| benefit5 | attend more appointments. | *0.578* | 0.199 | **0.676** | 0.485 | 0.488 |
| benefit18 | manage my other life responsibilities. | *0.425* | 0.333 | **0.590** | 0.543 | 0.431 |
| benefit17 | feel my time was being respected. | *0.393* | 0.330 | **0.556** | 0.524 | 0.391 |
| benefit9 | feel more in control of my pregnancy. | -0.230 | *0.873* | 0.202 | **0.759** | 0.616 |
| benefit11 | plan ahead and feel less stressed. | -0.002 | *0.815* | 0.401 | **0.814** | 0.663 |
| benefit7 | understand what was coming next in my pregnancy care. | 0.030 | *0.757* | 0.404 | **0.772** | 0.596 |
| benefit10 | feel confident that I was 'doing things right.' | 0.082 | *0.632* | 0.395 | **0.672** | 0.457 |
| benefit8 | know how to plan for tests. | 0.359 | *0.480* | 0.597 | **0.658** | 0.530 |
| benefit12 | explain my care plan to others (family, employers, other doctors). | 0.226 | *0.479* | 0.463 | **0.590** | 0.387 |
| benefit4 | understand how appointments differ. | 0.238 | *0.469* | 0.470 | **0.587** | 0.387 |
| benefit2 | be a stronger partner in my care. | 0.206 | *0.462* | 0.435 | **0.564** | 0.351 |
| benefit1 | increase my trust in the care team. | 0.258 | *0.461* | 0.487 | **0.589** | 0.397 |
| benefit3 | ask questions I wouldn't have thought to ask. | 0.341 | *0.379* | 0.528 | **0.547** | 0.387 |
| *Factor pattern displays item loadings. Italics indicate item factor loading.<br>**Factor structure displays correlations between individual items and the final two factors. Bolded correlations represent where items were interpreted as loading.<br>***Final communality estimates represent the proportion of each item's variance that can be explained by the factors. |  |  |  |  |  |  |

As an additional validation method, a three-factor EFA model with different rotation, an orthogonal rotation, was performed, with items loading similarly as the final three-factor model with oblique rotation, thus corroborating the final selected model approach. The three-factor model with orthogonal rotation validated the interpretation of the final selected model. See Figure 2 for the three-factor model results with orthogonal (varimax) rotation.

**Figure II.**
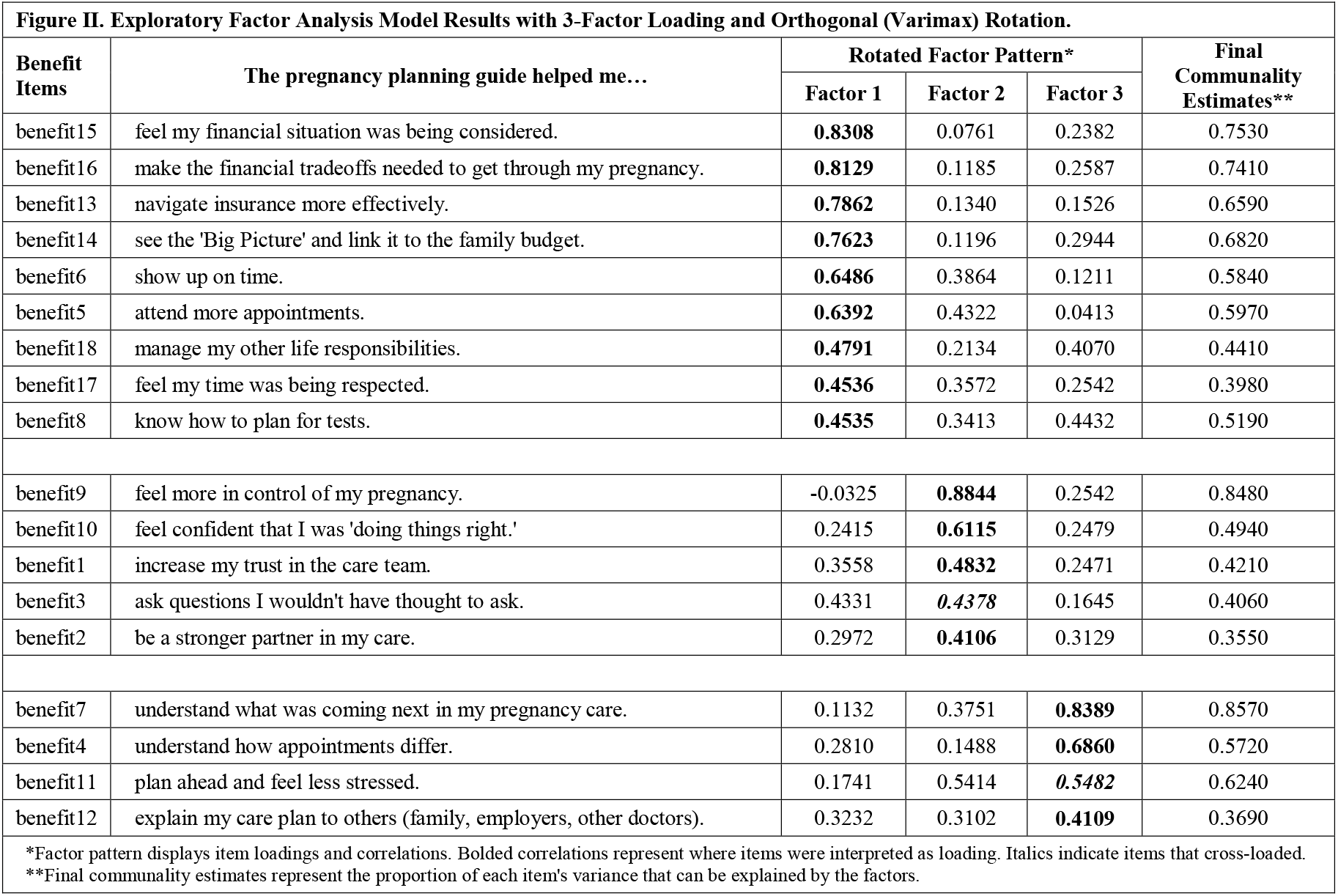
Exploratory Factor Analysis Model Results with 3-Factor Loading and Orthogonal (Varimax) Rotation.

## Discussion

The main purpose of this study was to identify key constructs underlying benefit items, or prenatal patients’ experiences, with the piloted cost tool. This study found three common underlying factors that encompassed the interrelationships between patients’ experiences as measured by the 18 benefit items: logistics, efficacy and [patient] understanding [of care plan]. EFA of the benefit items from prenatal patients who used the cost tool produced a three-factor model that is consistent with emerging themes from qualitative interviews of the mixed-methods study. The original goal of the cost tool was to act as a means to initiate cost conversations between patients and providers. A cost-related theme, labeled in this study as logistics, was a very clear and unwavering factor that emerged from the data. Throughout the study, other potential benefits of the pilot tool emerged: a means to know and plan for what will happen (understanding) and a means to reinforce one is doing things right, or to feel in control (efficacy). Interestingly, both of these themes were well-supported by the data as aligning with factors in this analysis.

Additionally, this study provides an analytic solution for mixed-methods studies. Specifically, this study excavated 18 benefits from in-depth interviews in a prior phase of this study using validated HCD methods (Erwin et al. 2019). The 18 individual benefits were then assessed among sample participants using both interview and survey data collection methods. After unifying the response option scales across data collection methods, this manuscript details the analysis process for identifying the common latent factors underlying all 18 individual benefits. One factor aligned clearly with the original intention of the tool, somewhat validating the construct validity of the target benefits. Furthermore, this analysis allowed for additional tool benefits of efficacy and understanding to be identified.

In addition to performing this factor analysis that identified three common factors underlying the 18 benefit items across the pilot study sample, it should be noted that this research team also examined the individual benefit items experienced due to use of the cost tool by income groups within the sample, defined by public or private insurance (Rivelli et al. 2022). Further, this team also detailed differences in cost-related conversations, specifically with whom sample prenatal participants were having them, and cost-specific benefits by income group due to use of the tool (Fitzpatrick et al. 2022).

The greatest utility of this analysis was that it grouped a large number of individual benefit items into three broader and more comprehensive constructs. With these broader constructs in mind as likely benefits, we will expand our pilot study to a larger group of prenatal patients and collect data using validated and standardized measures of the identified factors to further support the benefits of this cost tool. We hope to expand the use of this tool throughout multiple learning healthcare systems. This study will continue to explore the identified factors of logistics, efficacy and [patient] understanding [of care plan] as outcomes describing the utility of this cost tool. Furthermore, given so many items loaded on the logistics factor, separating items into two types of logistics – time and financial costs – will be considered. Then we will identify demographic- and pregnancy-related factors associated with prenatal patients’ experiences of these benefit “factors.”

### Strengths

This study provides a framework for unifying response options collected from a mixed-methods study in order to pursue quantitative analysis. This study also detailed a step-by-step analysis approach to identifying the underlying factor structure of the cost tool benefit items, as determined by prenatal patients’ benefit item responses. This study adds a quantitative component to better understand patients’ experiences with the cost tool as revealed through a qualitative human-centered design method (ie. card sort) and a survey alternative.

### Limitations

Because there is no “gold standard” tool or any other measure with which to compare and assess validity of these benefit items to measure tool utility, this current approach and the subsequent findings should be interpreted accordingly. Also, while ideal sample size for EFA is subjective and strict rules regarding sample size for EFA have mostly disappeared (Costello & Osborne 2005), a general consensus is between 3-10 participants per item or a reasonable absolute minimum sample size of 50 (De Winter, Dodou, Wieringa 2009). In our study, 71 participants responded to the 18 items, equating to a 3.94:1 ratio of participants per item, which is appropriate. Furthermore, results of this study confirmed strong data. Per Costello & Osborne (2005), the stronger the data, as defined by high communalities without many cross loadings plus several variables loading strongly (0.50 or better) onto each factor, the smaller the sample can be for accurate analysis.

## Data Availability

All data produced in the present study are available upon reasonable request to the authors

## Declarations

### Ethics approval and consent to participate

Prior to onset, study approval was granted by the health system’s institutional review board (#20-264E) and all patient informed consent was obtained in writing.

### Consent for publication

Not applicable

### Availability of data and materials

The datasets generated analyzed during the current study are not publicly available due to proprietary issues by the healthcare system but are available from the corresponding author on reasonable request.

### Authors’ contributions

AR analyzed and interpreted the factor analysis data. AR and VF were major contributors in manuscript writing. MS was a minor contributor in manuscript writing. VF and KE were responsible for the conception, design and securing of funding of this project. All authors read and approved the final manuscript.

## Acknowledgements

The authors would like to thank those who supported this study, specifically Andy Marek and Chris Blumberg in Research Analytics.

